# Methodology for tDCS integration with fMRI

**DOI:** 10.1101/19006288

**Authors:** Zeinab Esmaeilpour, A. Duke Shereen, Peyman Ghobadi-Azbari, Abhishek Datta, Adam J. Woods, Maria Ironside, Jacinta O’Shea, Ulrich Kirk, Marom Bikson, Hamed Ekhtiari

## Abstract

Integration of tDCS with fMRI holds promise for investigation the underlying mechanism of stimulation effect. There are 118 published tDCS studies (up to 1^st^ Oct 2018) that used fMRI as a proxy measure of neural activation to answer mechanistic, predictive, and localization questions about how brain activity is modulated by tDCS. FMRI can potentially contribute as: a measure of cognitive state-level variance in baseline brain activation before tDCS; inform the design of stimulation montages that aim to target functional networks during specific tasks; and act as an outcome measure of functional response to tDCS. In this systematic review we explore methodological parameter space of tDCS integration with fMRI. Existing tDCS-fMRI literature shows little replication across these permutations; few studies used comparable study designs. Here, we use a case study with both task and resting state fMRI before and after tDCS in a cross-over design to discuss methodological confounds. We further outline how computational models of current flow should be combined with imaging data to understand sources of variability in responsiveness. Through the case study, we demonstrate how modeling and imaging methodology can be integrated for individualized analysis. Finally, we discuss the importance of conducting tDCS-fMRI with stimulation equipment certified as safe to use inside the MR scanner, and of correcting for image artifacts caused by tDCS. tDCS-fMRI can address important questions on the functional mechanisms of tDCS action (e.g. target engagement) and has the potential to support enhancement of behavioral interventions, provided studies are designed rationally.

## 1. Introduction

Transcranial direct current stimulation (tDCS) applies direct current through electrodes placed on the scalp to modulate excitability (and consequently functioning) of the central nervous system (Y.-Z. Huang et al., 2017; Michael A Nitsche & Walter Paulus, 2000; Woods et al., 2016). In humans, tDCS is being used to investigate brain-behavior relationships and is currently under investigation for treatment potential in various neurological and psychiatric disorders (Andre Russowsky Brunoni et al., 2012). However, despite encouraging results in human neurophysiological experiments (M. A. Nitsche & W. Paulus, 2000; M. A. Nitsche & Paulus, 2001; Stagg & Nitsche, 2011), clinical trials (Antal et al., 2017; Marom Bikson et al., 2016; Boggio et al., 2008; A. R. Brunoni et al., 2013; Fregni et al., 2006; Lefaucheur et al., 2017; Valle et al., 2009) and detailed characterization of its physiological mechanisms of action in animal models (Marom Bikson et al., 2004; Ironside et al., 2018; Jackson et al., 2016; O’Shea & Revol, 2017), questions remain about protocol optimization, especially as relating to inter and intra individual variability (Chew, Ho, & Loo, 2015; Dyke, Kim, Jackson, & Jackson, 2016; Horvath, Vogrin, Carter, Cook, & Forte, 2016; Lopez-Alonso, Fernandez-Del-Olmo, Costantini, Gonzalez-Henriquez, & Cheeran, 2015; Worsching et al., 2017). The integration of tDCS with modern neuroimaging techniques (Saiote, Turi, Paulus, & Antal, 2013; Turi, Paulus, & Antal, 2012) is likely to represent a key methodological approach for advancing our understanding of the functional correlates of tDCS mechanisms in terms of changes in patterns of brain activation and for understanding individual differences in response to stimulation (Buch et al., 2017; Esmaeilpour et al., 2018; Giordano et al., 2017).

Magnetic resonance imaging (MRI) supports noninvasive imaging of brain structure and function. The former is capable of providing high spatial resolution in terms of identifying anatomical regions while the latter allows the study of dynamic physiological changes (Symms, Jäger, Schmierer, & Yousry, 2006). Functional MRI signal is approximately proportional to a measure of local neural activation integrated over a spatial extent of several millimeters (Heeger & Ress, 2002). Integration of tDCS with MR imaging techniques provides a tool to directly perturb neuronal function while monitoring brain state. Therefore, it enables researchers to study not only how stimulation modulates targeted brain regions, but also how tDCS modulates activity across the brain in the context of anatomical and functional connectivity. In addition, this integration may also provide critical insight for understanding how, where and when stimulation is likely to be most effective – useful for optimization purposes.

In recent years, there has been increasing interest in functional magnetic resonance imaging (MRI) techniques to study the effects of tDCS - both in healthy controls and clinical populations. The first paper that utilized fMRI to investigate how tDCS modulates neuronal activity was published in 2001, where imaging was done sequentially before and after stimulation (Baudewig, Nitsche, Paulus, & Frahm, 2001). The majority of studies (>80%) to date have used fMRI to evaluate effects of stimulation on cortical regions underneath the electrodes as well as distal areas. However, it also can potentially help to optimize stimulation protocols or may serve as a possible biomarker for variability in responsiveness among subjects (Cavaliere et al., 2016; Kasahara, Tanaka, Hanakawa, Senoo, & Honda, 2013).

The organization of this document centers around methodological aspects of tDCS integration with MR imaging techniques. The goal of this document is not to recommend, embargo or critique a single paper. While we review the overall literature to assess the breath of technology and highlight the diversity (lack of consistency in methodological approach), individual papers are cited applicable to specific arguments. We start with a literature review to analyze the parameter space for trial design in tDCS-MR imaging studies and discuss how MR imaging could contribute in tDCS montage selection, response prediction and as an outcome measure in terms of neural activation. Through a case study we then introduce potential confounds in the experimental design of tDCS-fMRI studies; where best practices from fMRI trials and tDCS trials (each in isolation) nonetheless produce unexpected interactions in combined tDCS-fMRI experiments. We further discuss practical considerations regarding MR-compatible stimulation devices and imaging artifacts. Lastly, we discuss how computational finite element models could be integrated in MR imaging studies to reduce variability in responsiveness. Here again we use a case analysis to demonstrate how modeling and image processing can influence outcomes.

## 2. Literature review of parameter space in tDCS-MR imaging trial design

The authors adopted a systematic search strategy in accordance with the most updated preferred reporting items for systematic reviews and meta-analyses (PRISMA) (Moher, Liberati, Tetzlaff, & Altman, 2010). Electronic search was performed in PubMed using logical combinations of the following keywords: (“fMRI” OR “functional MRI” OR “functional magnetic resonance imaging” OR “rsfMRI” OR “resting-state MRI” OR “fcMRI” OR “functional connectivity MRI”) AND (“tDCS” OR “transcranial direct current stimulation”). The PubMed research database was searched from inception to 1st October 2018 with restriction on human studies in English.

Original studies of functional MRI with different paradigms for combining with tDCS were included in this review. We excluded reviews, guidelines, book chapters, case reports, clinical trial study protocol/design articles, studies on the combination of fMRI with non-tDCS techniques (e.g., TMS, tACS, tPCS, and tRNS), studies on combining tDCS with electrophysiological/neuroimaging methods other than fMRI and ASL (e.g. EEG, ERP, MEG, DTI/DWI, MRS, and structural MRI), and tDCS or fMRI-only articles (Fig 1).

**Figure 1:**
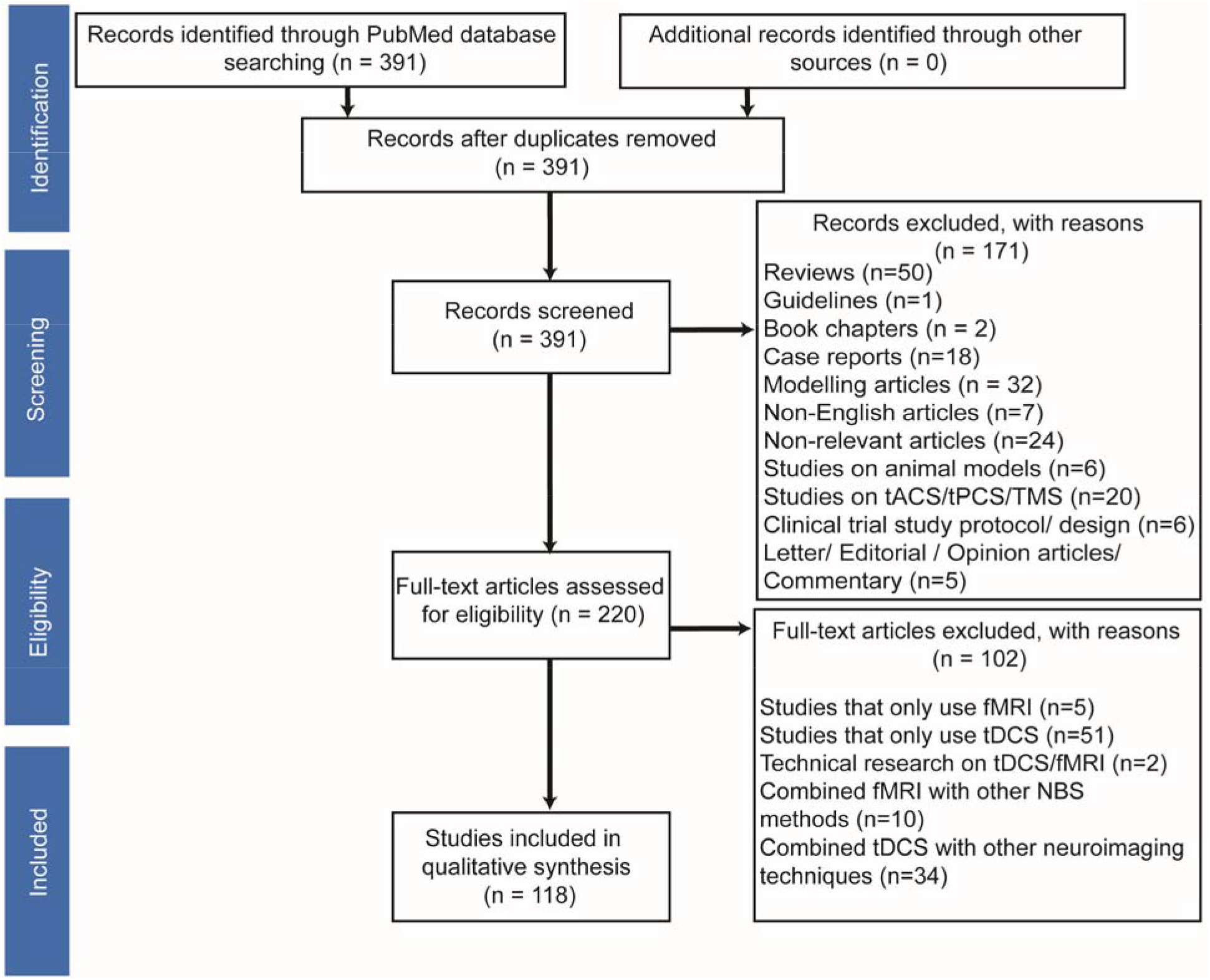
Search strategy for inclusion, exclusion criteria. To identify relevant studies, we searched PubMed using logical combination of keywords “tDCS” OR “transcranial direct current stimulation”) AND (“functional magnetic resonance imaging” OR “fMRI” OR “functional MRI” OR “fcMRI” OR “functional connectivity MRI” OR “rsfMRI” OR “resting-state MRI”.

The titles and abstracts of the 391 full-length articles found by the search were screened. 171 studies were excluded based on the above exclusion criteria to yield 220 full-length articles. Then, the same investigators scrutinized the full texts of relevant articles (n=220) against the above inclusion and exclusion criteria, which resulted in further exclusion of 102 articles. This resulted in 118 articles included in the qualitative synthesis (Fig 1).

## 3. tDCS-MR imaging: trial design parameter space

The study protocol is selected based on several assumptions and hypotheses such as time course of tDCS effect (Michael A Nitsche & Walter Paulus, 2000), cellular mechanism of stimulation (Marom Bikson et al., 2004; Rahman et al., 2013), state dependency of tDCS as a subthreshold stimulation method (Hsu, Juan, & Tseng, 2016; Li et al., 2019) and dose-response relationship. Ultimately, these considerations get to the heart of how tDCS research benefits from functional MR imaging and how to address inherent challenges in experimental design. Parameter space in tDCS-fMRI trial design includes:

### 1) fMRI timing relative to tDCS

In research studies timing can be divided into three approaches*-<u>Sequential-inside scanner</u>* <u>approach</u>, where imaging data are collected before and/or after tDCS intervention inside the bore of magnet, *<u>sequential-outside scanner</u>* <u>approach</u>, where imaging data are collected before and/or after tDCS intervention with stimulation done outside the scanner and *<u>concurrent</u>* <u>approach</u>, where imaging data are acquired during (and often before and after) tDCS stimulation inside the scanner. The sequential approach allows evaluation of the functional brain state before stimulation and/or the lasting “after-effects” of stimulation, while the concurrent approach allows studying the effects of stimulation applied during ongoing neural activity. The two main difference between “sequential-inside scanner” and “sequential-outside scanner” approach is tDCS timing and posture during stimulation (i.e. upright position vs supine). Modeling studies suggest that inter-postural difference influences cerebrospinal fluid (CSF) distribution and consequently current flow in brain (Mikkonen & Laakso, 2019).

Prior to MR-compatible tDCS systems, studies were limited to the sequential-outside scanner approach, which represents the majority of studies in this review (72% of studies). In longitudinal studies, there can be follow-up imaging sessions hours or days after stimulation or several stimulation sessions with imaging over the course of days or weeks (Fig 2).

**Figure 2:**
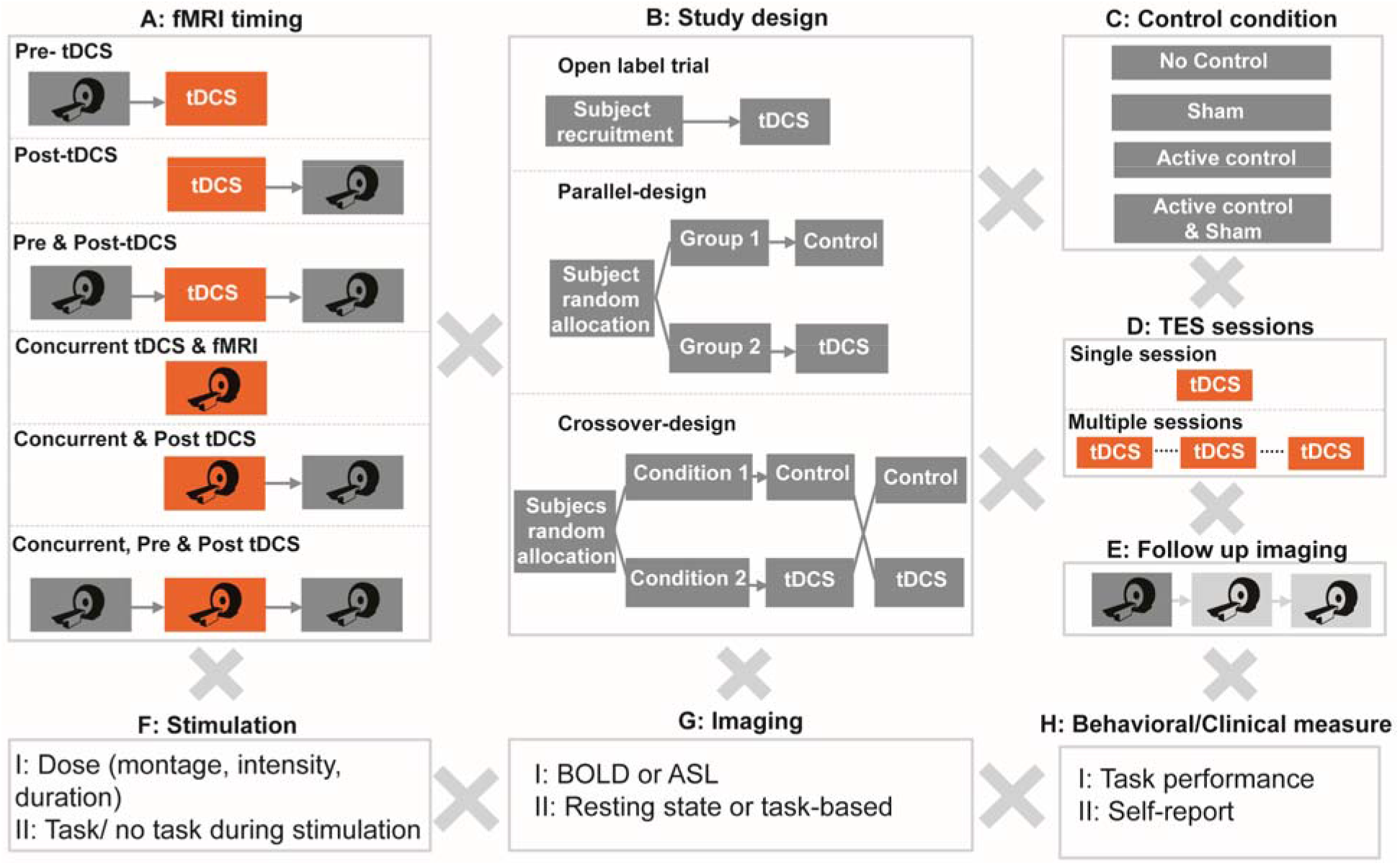
Permutations in trial design for integration of MRI-based neuroimaging methods with tDCS. (A) fMRI timing can occur pre and/or post-stimulation outside scanner (sequential-outside scanner approach), pre and/or post-stimulation inside scanner (sequential-inside scanner approach) or concurrently during stimulation to evaluate tDCS effect on ongoing neural activities. (B) Study designs include single arm open label studies with no control condition, parallel approach where subjects are randomly assigned to either active or sham group and crossover approach, where subjects participate in both active and sham sessions with a random order. (C) Control conditions consist of sham, active control or both. (D) tDCS may be applied over multiple sessions with an expectation of cumulative (time) effect. (E) Imaging may include multiple follow up sessions to evaluate after effects longitudinally. (F) Stimulation dose (i.e. electrode montage, stimulation duration and intensity) and combination with task will determine outcomes. (G) Essentially any functional imaging sequence can be utilized to evaluate stimulation effects depending on the study questions/hypothesis. (H) tDCS-imaging study designs can combine a multitude of other objective or subjective assessments concurrent or combined with imaging and/or tDCS.

### 2) MR imaging protocol and sequence (task/ rest and BOLD, ASL)

Stimulation could be paired with specific tasks to investigate the effect of stimulation on ongoing task dependent activity/connectivity (i.e. functional targeting) or be done during resting-state (Fig 3). There are different perfusion fMRI sequences available, like arterial spin labeling (ASL). However, the majority of tDCS-MR imaging studies to date have used task and resting-state BOLD (blood oxygenation level dependent) fMRI (95% of studies). There is a wide range of specific MRI sequences for ASL and fMRI as well as a large variety of post-processing methods that may also lead to different results (Wörsching et al., 2016). Therefore, care must be taken to ensure the experimental design and methods chosen are robust and sensitive to capturing tDCS effects.

**Figure 3:**
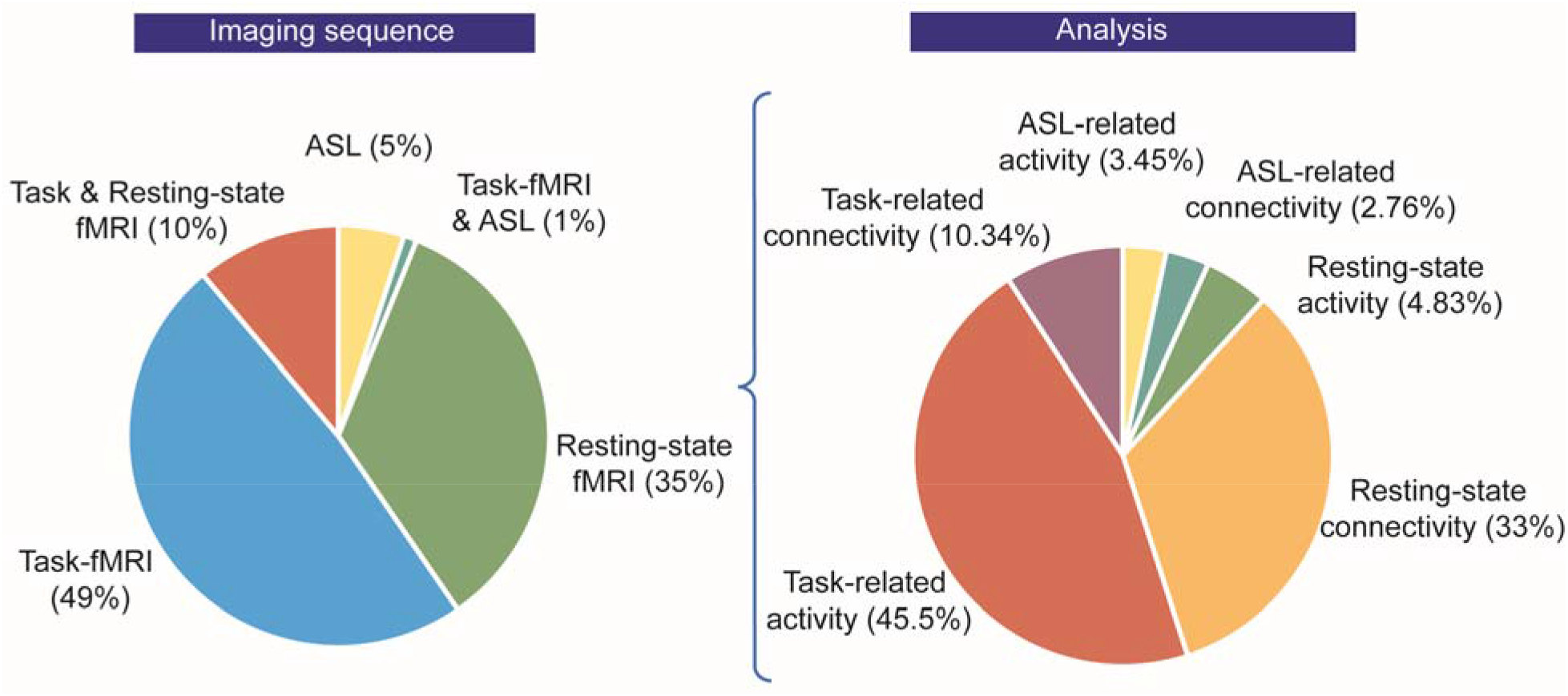
Functional imaging sequences and analysis methods used in literature of tDCS-fMRI studies.

### 3) Control condition (sham and/or active control)

The usual approach for blinding a study is to use a sham stimulation (no stimulation during the session but inducing sensations at the beginning and/or end of the session via ramp up/ ramp down of current). However, inconsistency in sham-controlled studies in tDCS literature resulted in debates on whether different sham protocols are equivalent (Fonteneau et al., 2019). Another approach in blinding a study is active control, which refers to a condition where the stimulation is performed over an area irrelevant for the purpose of study.

### 4) Study design (parallel, crossover or open label)

The majority of tDCS-fMRI studies are designed either as randomized controlled trials (89%) which consist of “parallel studies” (31%) where participants are randomly assigned to either an active or sham tDCS group, and “cross-over studies” (58%), where each participant is assigned to both active and sham sessions in a randomized order (Fig 2 B). However, “open label studies” are used as well, depending on study objectives (e.g. intent-to-treat, dose optimization) and only represent 11% of studies.

### 5) Stimulation dose and individual factors

Dose can be defined base on several factors such as montage, electrode size, duration of stimulation, current intensity and waveform (see (Peterchev et al., 2012) for detailed information). Anatomical and biological factors such as head size (Marom Bikson, Rahman, & Datta, 2012), skull thickness and individual gyrification of cortex (Opitz, Paulus, Will, Antunes, & Thielscher, 2015), structural and functional connections (C. Rosso et al., 2014) neurotransmitter levels (Krause & Cohen Kadosh, 2014), age (Minhas, Bikson, Woods, Rosen, & Kessler, 2012) and genetics (Thair, Holloway, Newport, & Smith, 2017) are among the parameters that can cause variability in response to a given dose across participants. Notably, models and measures of current flow show that for the same applied current, substantial differences in induced electric field in brain can occur across individuals (Esmaeilpour et al., 2018; Truong et al., 2014).

### 6) Objective (cognitive, clinical or behavioral) or subjective (clinician rated or self-report) measurements

Studies can use objective and/or subjective measures before, during or after stimulation. The measure should be selected in harmony with other parameters of the study to enhance the probability of detecting an effect and reducing noise.

When behavioral/clinical outcome measures are collected these can be correlated with functional imaging measures. In principle, the correlation between changes in brain activation with changes in behavioral/clinical outcomes would provide evidence for target engagement; namely providing support that the behavioral/clinical outcome is associated with neuromodulation of a functional brain regions or networks. However, the numerous permutations in study design (Fig 2) including imaging sequence (Fig 2 G) and known dependence of tDCS effect on dose (Fig 2 F), time, and brain-state (task), as well as questions on ideal trial design and sham (Fig 2 C) provides much complexity. At a minimum, investigators must be cognizant of experimental limitations and carefully design suitable experiments to address the specific research question, as unexpected interactions may produce misleading study conclusions. For example, it may be important to consider the differences between diffuse stimulation when conventional two-pads as opposed to focal stimulation with HD-tDCS (Datta et al., 2009b). For these reasons of potential heterogeneity, care must also be taken in any attempt to extrapolate across tDCS-imaging trials or meta-analysis (M. Nitsche, Bikson, & Bestmann, 2015). Moreover, we show next that tDCS-imaging studies must be designed with nuance on both stimulation and MRI parameters, such that combining a trial design optimized for stand-alone imaging may not be suitable when integrating tDCS, and vice versa.

### 7) Task/ brain state

Task-based fMRI is used to inform our understanding of how tDCS modulates brain activity while performing a task. In study design, it is important to consider that tDCS effects are task-specific; meaning stimulation does not necessarily produce the same effect while performing two different tasks (Saucedo Marquez, Zhang, Swinnen, Meesen, & Wenderoth, 2013). In contrast, resting-state fMRI is a technique performed in wakefulness without a superimposed task to execute. If the nature of tDCS is (profoundly) influenced by brain state (Li et al., 2019; A. Shahbabaie et al., 2014), stimulation of different individuals with same current flow profile could lead to different results based on their brain state.

### 3.1. Case study for nuance in trial design

We describe an fMRI study design which adapted best practices for imaging without stimulation and integrated a typical tDCS stimulation trial design (typical in stand-alone studies with imaging), yet resulted in complex interactions (Alireza Shahbabaie et al., 2018). In a double blind, crossover (counterbalanced order), sham controlled study on methamphetamine abstinent subjects (Fig 4), functional MR scans were acquired using sequential-outside scanner approach and subjects completed a cue-induced craving task inside scanner (a block-design task with two types of blocks: the active blocks consisted of 4 drug-related images and the control block consisted of 4 neutral images matched for psychophysical features with drug-related images). For each scan, response to drug-related blocks (cue induced craving) was contrasted with neutral. Subjects also rated their immediate methamphetamine craving before and after each session of stimulation and imaging (i.e. 4 timepoints) (Fig 4, A). There was at least a one-week washout period between the two sessions. Importantly, in this design, drug cues presented during imaging served as a stimulus to activate brain regions associated with craving. The induced craving was measured with fMRI and subjective reports (i.e. self-reported visual analogue scale pre and post-imaging).

**Figure 4:**
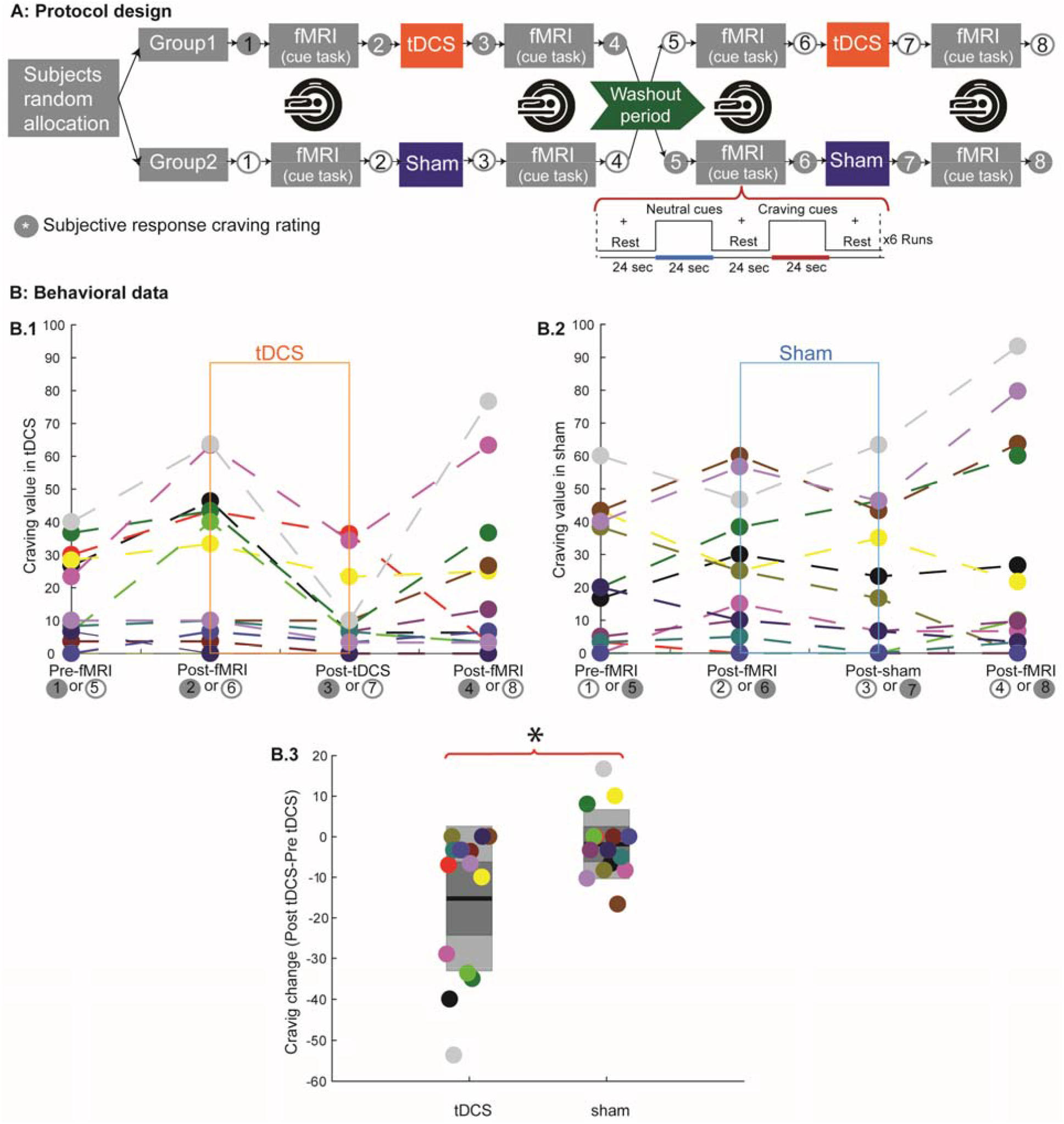
An experiment design and self-report (subjective) results in a double blind, crossover, sham controlled study. (A) Each subject underwent a counterbalanced study design with two stimulation types (real and sham) on two separate days with a one-week wash out period. Functional MRI (fMRI) data were acquired before and after each stimulation session. Subjects rated their immediate methamphetamine craving before and after both stimulation and imaging sessions on a visual analogue scale (VAS), with a score range from 0 to 100, where 0 and 100 indicated ‘no craving’ and ‘extreme craving’, respectively. (B) Self-report results for 4 different timepoints within the experiment. Subjects rated their craving level before and after each functional MR scan and stimulation session (i.e. sham, active). (B.3) tDCS significantly reduced craving compared to sham (p<0.05). (B.4). Study includes 15 subjects (colored circles represent each subject).

The key questions were 1) whether tDCS reduced drug craving (self-reported measure) heightened by drug-cues presented before tDCS/sham, and was this benefit maintained when second set of drug-cues were presented, 2) does tDCS compared to sham changed brain activation in regions associated with craving. Notwithstanding this sophisticated design for imaging trials, in this case tDCS produced a significant change in self-reported craving (Wilcoxon test, mean score change in tDCS session= −15.42 ± 5.42 *SE*, mean score change in sham session = −1 ± 2.63 *SE*; *p* = .03). This means subjects in the tDCS arm had less self-reported craving after stimulation compared to sham condition (before the second fMRI, Fig 5, B). This effect could be due to the non-linear interaction of tDCS effect with individual’s brain state (i.e. activations remained after the first cue-induced craving task).

**Figure 5:**
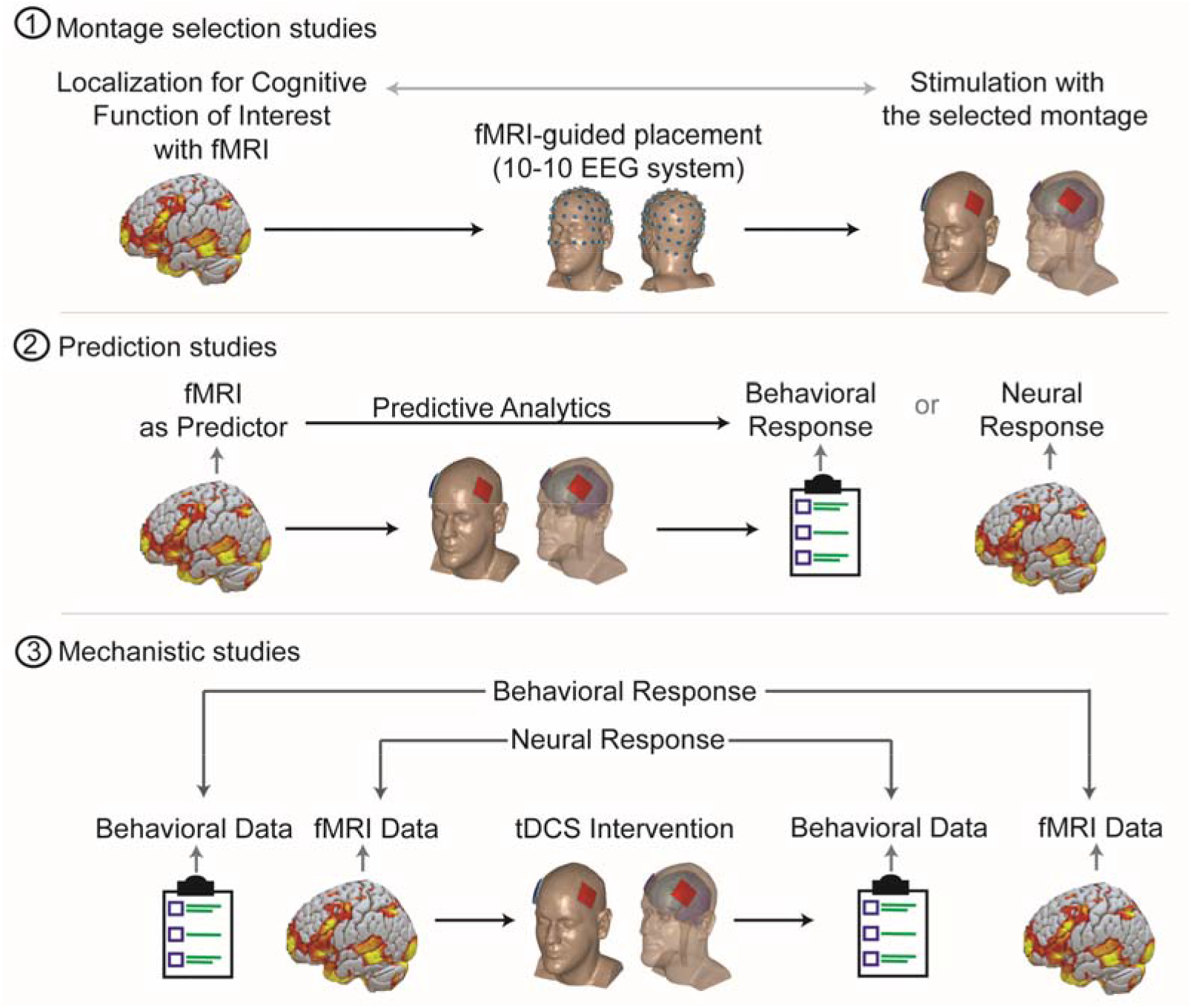
Three major roles functional MRI can play in integration with tDCS. 1) Montage selection studies: functional imaging could contribute to montage selection to localize stimulation to target functionally activated brain areas. (2) Prediction studies: baseline fMRI could provide baseline measures of neural network activation and inform the interpretation of later behavioral/neural responses to tDCS (i.e. “Baseline fMRI as Predictor”). (3) Mechanistic studies: utilizing fMRI to investigate brain changes underlying tDCS effects and ultimately determine where, when and how stimulation affects brain function and associated behavior.

In analyzing neural activation changes induced by tDCS, three nested fixed effect comparisons are required for each subject: at the first level of analysis, the contrast of cues (drug vs. neutral images) in the first and second fMRI scans in each session (time effect); and then at the second level the contrast across conditions (tDCS vs. sham). At the third level, these contrasts are then passed up to test for a group effect across participants using a mixed effects analysis.

In addition to non-linear dependence of tDCS effect on brain state, second exposure of a task that involves emotion/motivation (i.e. first drug-cue task may have residual effect that interact with second exposure) would make non-linear interaction with tDCS effect more complex.

For a tDCS trial focused only on determining the subjective (self-report) effects of tDCS on baseline craving, this protocol might not optimal - even disregarding the cost/burden of fMRI. Alternatives for an imaging study would be 1) a straightforward design for acute changes that uses resting-state imaging before and/or after stimulation, 2) task-based fMRI only after tDCS/sham, 3) tDCS/sham over multiple sessions followed by assessment of drug craving, as well as other outcome measures significant for addiction medicine such as abstinence. Each of the above options for integrations of task based or rest fMRI with tDCS will have their own pros and cons. The goal of this case analysis is not to question the validity of any given tDCS-imaging trial, but to say that although there are validated and thorough imaging trial designs and accepted protocols to test behavioral or neurophysiological effects of tDCS, their integration with fMRI is not trivial. Investigators should be cognizant of all the limitations and possible interactions of the design.

## 4. How tDCS studies can benefit from functional MR imaging

Based on the literature review, functional MR imaging is being used to benefit tDCS studies in three different ways: study design, outcome evaluation at the neural network level, and serving as potential biomarkers for responsiveness to tDCS. When functional imaging is conducted before stimulation, it could potentially provide valuable information about functional localization to guide montage selection (see montage selection, Fig 5). Functional imaging before the tDCS intervention could also be used to quantify factors predictive of response to tDCS (see prediction studies, Fig 5). Imaging the brain response to tDCS during or before and/or after stimulation is used to reveal the immediate effects of ongoing tDCS on functional brain activation and/or the lasting after-effects of stimulation (see mechanistic studies, Fig 5).

### 4.1. Montage selection using MR imaging

An essential question in tDCS protocol design is which stimulation montage should be selected to 1) optimally target the brain area to modulate– broadly called anatomical targeting; 2) understand which brain regions are active in a specific task/person and whether targeting active regions increases stimulation efficacy – broadly called functional targeting (Guleyupoglu, Schestatsky, Edwards, Fregni, & Bikson, 2013). MR imaging could provide an index of neural activation and is therefore invaluable for functional targeting purposes. Optimal targeting of a selected brain region requires head modeling based on structural MRI (Datta et al., 2009a; Miranda, 2013). In the literature, 11% of studies (13 out of 118) utilized task or resting MRI for designing the tDCS montage (Baker, Rorden, & Fridriksson, 2010; Clark et al., 2012; Hu et al., 2017; Mizuguchi, Uehara, Hirose, Yamamoto, & Naito, 2016; Woods et al., 2014). For instance, Fischer and colleagues selected a stimulation montage to target a motor network identified by resting-state functional connectivity. The hypothesis was that stimulating motor network would increase tDCS effect compared to motor area (M1). Using a modeling approach, 8 HD (high definition) electrodes were used in an arrangement to target the defined motor network with the purpose of increasing stimulation efficacy (Fischer et al., 2017a).

### 4.2. Prediction studies

A challenge in tDCS studies is inconsistency in outcome; the effects of tDCS appear to vary among individuals (Datta, Truong, Minhas, Parra, & Bikson, 2012; Krause & Kadosh, 2014). However, the sources of variability remain to be determined. Functional imaging may provide critical information for response prediction (Fig 5). In the literature, there are 4 studies that utilized functional or resting-state MR imaging as a predictive measure of responsiveness (Cavaliere et al., 2016; Kasahara et al., 2013; Polania, Paulus, & Nitsche, 2012; Charlotte Rosso et al., 2014). For example, in minimally conscious state (MCS) patients, resting state functional connectivity before tDCS was suggested as a potential biomarker of responsiveness, detecting neural conditions necessary for tDCS to facilitate transitory recovery of consciousness and potentially improve behavior in MCS (Cavaliere et al., 2016).

### 4.3. Mechanistic studies

The majority of studies (101 out of 118) in this review reside in this category. In this group of studies, MR-based functional imaging methods (i.e. ASL, task and resting-state) are used to investigate underlying neural correlates of tDCS mechanisms in terms of changes in brain activation not only in targeted areas but across functionally activated task-relevant brain regions and large-scale neural networks (Keeser et al., 2011; M. Meinzer et al., 2012; Peña-Gómez et al., 2012; Wörsching et al., 2016) (Fig 5).

## 5. Practical considerations when combining tDCS with functional MRI

Concurrent tDCS/fMRI data acquisition as well as sequential-inside scanner experiments have additional considerations over and above those required for sequential-outside scanner tDCS and fMRI experiments. These include: MRI-compatible stimulator setup, choice of RF coil, electrode hydration, stray fields induced from tDCS wires, and disambiguating physiological signal from tDCS artifacts in the fMRI data.

### 5.1. Safety

TDCS in general is considered safe and well-tolerated (M. Bikson et al., 2016). A safety concern in combining tDCS and MRI is the possibility of heating under the electrodes due to radio-frequency pulses of scanner (Saiote et al., 2013). MRI compatible tDCS requires careful insertion of specially designed MRI-compatible tDCS cables and conductive polymer electrodes through the magnet through waveguide or filter box and into the magnet bore. Outside the MRI, saline-saturated sponges are typical in tDCS while gel is used for HD-tDCS, but for a range of reasons (including increased properties for electrolyte spread during prone subject adjustment, drying over extended time), conductive paste is preferred in the MRI. Conductive paste, such as ten20 paste, should be applied between the MRI safe conductive polymer electrodes and the scalp. The cables leading to these electrodes should be in series with RF filters and run parallel to the bore, without loops and away from the subject to prevent the risk of eddy current induction and potential RF burns. In general risk assessment should rely on overall protocol and imaging sequence rather than the device in isolation. This is of particular importance as newer imaging sequence and hardware, for example multiband sequences and stronger/faster gradients, become available.

### 5.2. MRI-compatible stimulator setup

A main hardware precaution to be aware of when bringing any electronic equipment into the MRI scanner is not allowing the wires to become RF transmitters and introduce noise into the MRI data. To prevent this, currently two similar hardware configurations for MRI-compatible tDCS are used (Marcus Meinzer et al., 2014; Williams et al., 2017). The procedures are described below:

I. Connect the leads from the tDCS power supply to an RF filter box (MRI control room), from the RF filter box to a nonmagnetic, shielded local network cable that enters the MRI scanner room via the RF waveguide tube, carefully oriented to reduce resonant capacitive coupling, and then finally sent through another RF filter box (MRI-compatible, inside scanner room) before connecting to the tDCS electrodes (Fig 6, A).

**Figure 6:**
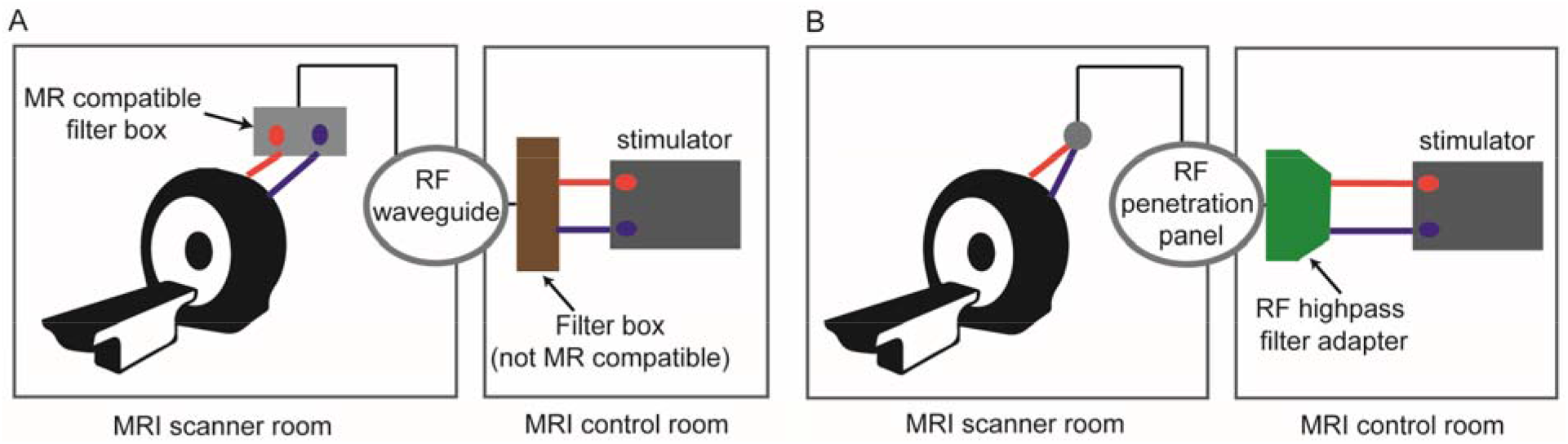
Different MR-compatible stimulator setups inside scanner. (A) setup comprises of two filter boxes (inside scanner room (MR-compatible) and outside scanner room (not MR-compatible)) and cables come from the control room through RF waveguide tube. (B) setup include a RF filter adapter that connects to RF penetration panel (device will have the same ground as the RF shield of MRI room).

II. Connect the leads from the tDCS power supply (MRI control room) to an RF filter attached to the penetration panel, and then connect the other end of the penetration panel (MRI scanner room) to the electrode leads (Fig 6, B).

Ideally, better imaging quality is expected when the tDCS wiring is run through the RF penetration panel as this further reduces the possibility of introducing electromagnetic noise picked up by the receiver MRI coils. While setup II inherently accomplishes this by having the RF filter mount directly onto the penetration panel, setup I can be modified slightly to include extra connections to send the wiring through the penetration panel rather than the waveguide and is the recommended approach.

### 5.3. RF coils

Many fMRI experiments utilize receive-only parallel RF coils (16, 32, or 64 channel head coils) to detect signal. However, as the number of coils, and therefore sensitivity increases, the empty space inside the head coil typically decreases. MRI compatible headphones and head padding further restrict available space, making some montages especially challenging. Accommodating particular montages may require advanced experimental consideration. In many cases the simplest solution is to compromise by using a larger, but less sensitive head coil.

### 5.4. Electrode preparation

In addition to these MRI-specific hardware modifications, it is important to be aware of modifications to the electrode setup. Saline soaked sponges have disadvantages in imaging settings. They tend to dehydrate over the course of the experiment. One solution, developed for a PET study (DosSantos et al., 2012), is to use two long cannulas in each sponge connected to syringes with saline slowly supplemented over the course of the stimulation/scan. Use of saline is also complicated because fluid may be displaced from the sponges and potentially bridge the electrodes and/or create loops, compromising both the integrity of the experiment and the safety of the subject. The recommended modification is to use electrically conductive pastes, but special precautions should be taken (Woods et al., 2016). With conventional (pad) tDCS, penetration of the conductive rubber electrode through the paste and contacting skin will produce skin burns, hence it is important to ensure minimum recommended paste thickness of ∼3 (preferably ∼5 mm) (Woods et al., 2016). With HD-tDCS, the plastic holder controls this separation. While using these pastes prevents dehydration, it is still possible that the paste will create bridges between electrodes if electrodes are placed too close to one another or physically shifted as part of subject/coil adjustment in the scanner (Fig 7).

**Figure 7:**
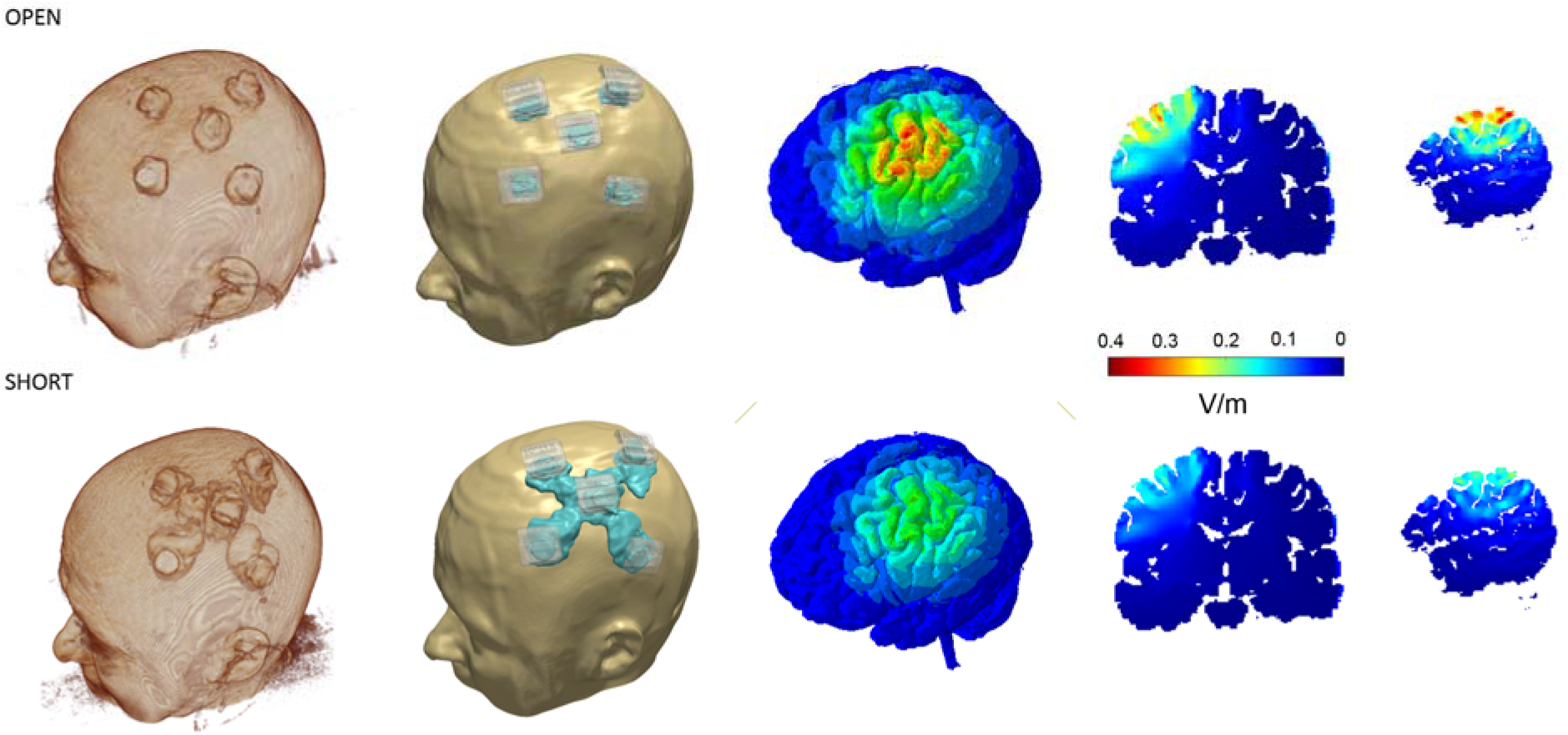
Induced cortical electric field magnitude for the montages considered. From left to right: 3D surfaces of skin and brain, cross-sectional coronal and sagittal images. OPEN: Enough gel for satisfactory impedance in the interface between electrode and skin but gel is contained in HD cups. SHORT: Excessive gel smeared between adjacent electrodes reflecting excessive gel application and/or head-gear motion. This results in highly conductive path for current to flow above the surface of skin (“short circuit”) that reduces current penetration to the brain.

### 5.5. Voltage compliance

MR safe electrodes typically have higher resistance than conventional tDCS electrodes. Therefore, some standard tDCS devices are not capable of providing sufficient voltage to induce typically used experimental current amplitudes (e.g. 2 mA) whilst using these electrodes. For example, if voltage compliance of a device is 40 volts and it needs to provide 2 mA current for stimulation, the resistance should be less than 20 KΩ (i.e. R=V/I) otherwise the device could not provide the target 2 mA for intensity. Comprehensive test of stimulation device is required before starting any experiment regardless of device brand or type.

### 5.6. Stray magnetic field from tDCS wires

The interaction of tDCS and MRI equipment creates other additional potential complications. For example, despite isolating the tDCS wires to filter out potential RF noise, simply passing current through the wires and electrodes induces stray magnetic fields that will increase magnetic field inhomogeneities and potential artifacts (Fig 8). Because the current density in the wires is generally much larger than that flowing through the brain, the induced local magnetic field from the tDCS wires is also much larger than that from currents in the brain. The resulting inhomogeneities can be difficult for field shimming to correct, which may influence fMRI adversely. While it is possible to calculate and subtract these stray fields, in practice this is challenging and rarely done (Göksu et al., 2018).

**Figure 8:**
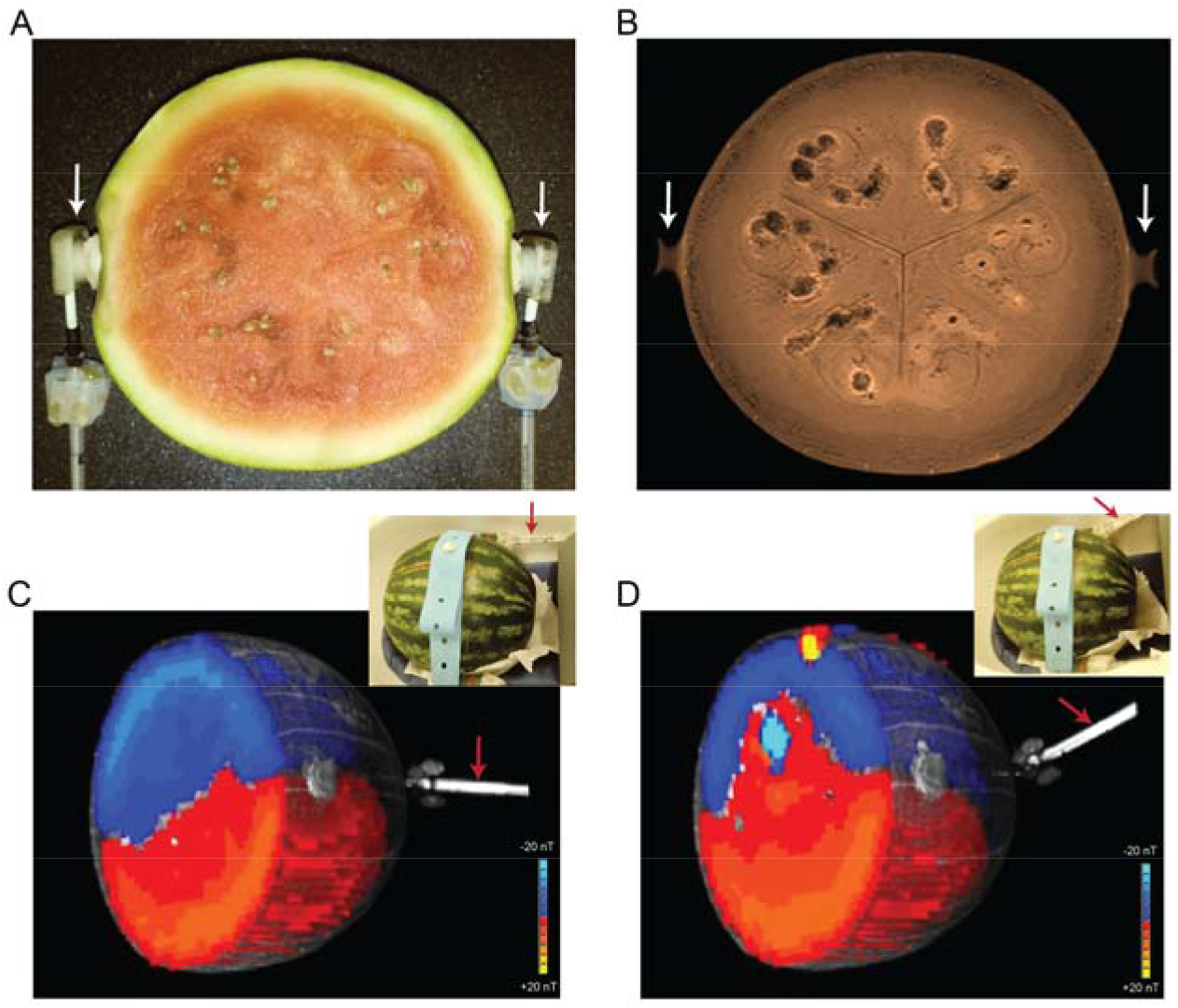
Effect of current induced stray magnet field on Bz in watermelon. Electrodes (white arrows) are positioned on left and right side along the equator (orientation visible in proton density MRI in (b)). MGE data from +/-2mA current injection was used to calculate effective Bz(+4mA) for cables oriented along z (c) and ∼25° from z (d). Cables immersed in agar (red arrows) provide contrast in the T1 underlays of the Bz images (c), (d). After experiments, the watermelon was sliced and photographed (a) at the level of (b), showing the structural sources of heterogeneity in (b).

### 5.7. Data quality considerations for concurrent MR imaging & tDCS

In the concurrent tDCS/fMRI approach, electrical current interacts with the magnetic field generated by the scanner, and therefore can potentially result in warping of the images acquired. This artefact must be considered as it can potentially cause false positive changes. The magnitude and nature of any artefacts is likely to depend on the exact experimental setup. When stimulation is performed inside scanner, effect of stimulation should be considered in image quality (i.e. reducing signal-to-noise ratio (SNR)) (Saiote et al., 2013).

One study using functional imaging and head models in two cadavers strongly suggested that tDCS could potentially cause significant BOLD signal changes (Antal et al., 2014). Another study demonstrated susceptibility artefacts underneath the electrodes restricted to skull layer and no visual evidence of any distortion in brain EPI images (Antal, Polania, Schmidt-Samoa, Dechent, & Paulus, 2011; Gbadeyan, Steinhauser, McMahon, & Meinzer, 2016). Therefore, careful inspection of signal in concurrent data acquisition protocols is of critical importance to diminish concerns over false positive results.

Taking precautions to mitigate the above negative effects of introducing tDCS concurrent with fMRI acquisition may not fully negate the potential for poor data or erroneous results, however. It is noteworthy that often the region most likely affected by tDCS induced artifacts is the same as that from which an experiment tests for changes in tDCS-driven functional activation—i.e. the region under the anode/cathode (Fig 9). In a concurrent tDCS and fMRI experiment (Shereen et al, in preparation), false positive fMRI results persisted even when comparing finger tapping to rest condition within the tDCS stimulation session inside scanner (Fig 9). This increases the likelihood of false positives (Antal et al., 2014) and it is difficult to disambiguate the extent to which increased fMRI BOLD results are due to genuine tDCS-induced neuromodulation of brain activity rather than tDCS-induced false positive artifacts. Moreover, traditional controls (sham, active sham at different location) would not account for dose specific artifacts.

**Figure 9:**
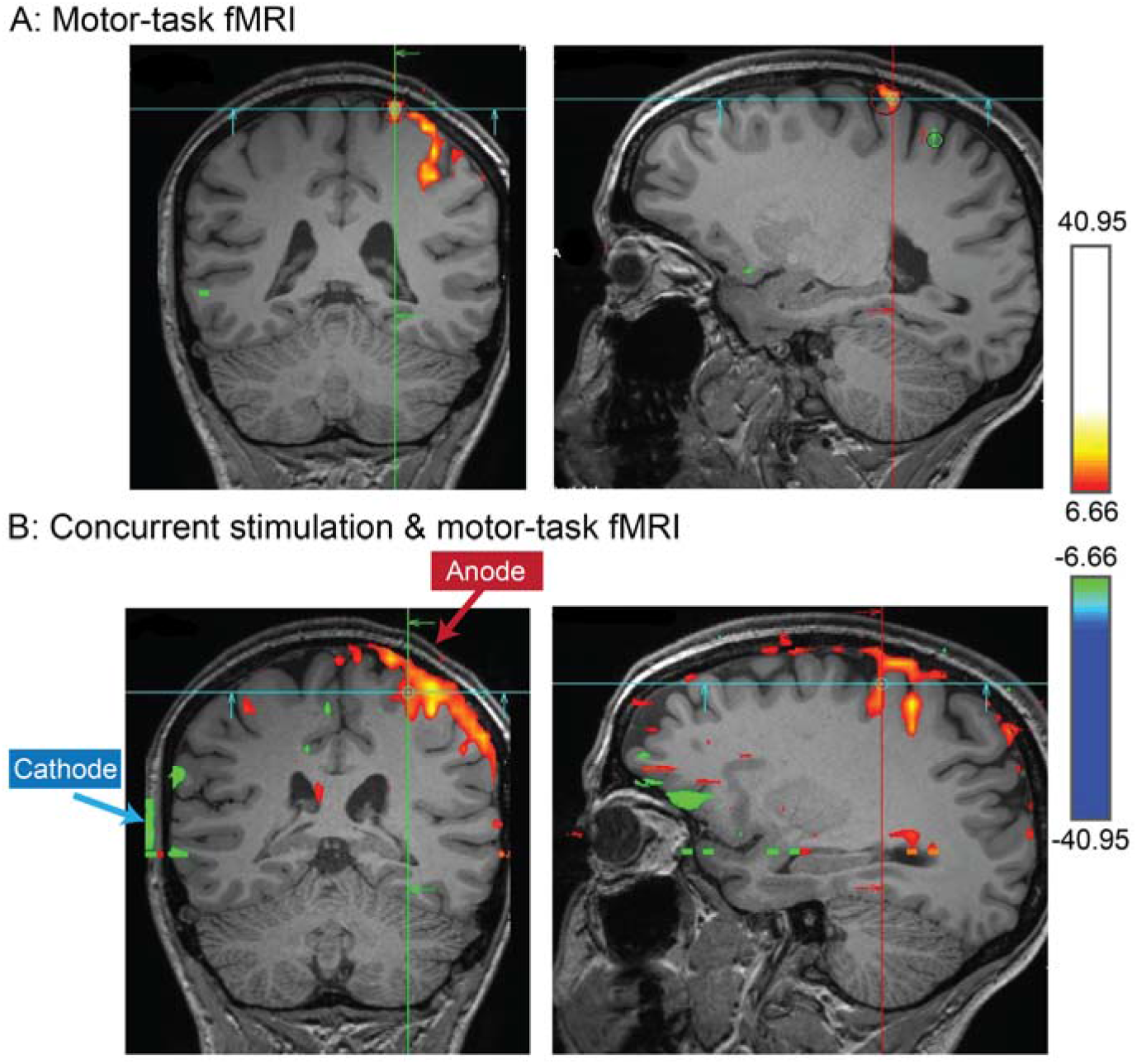
Concurrent stimulation & imaging artifact. fMRI activation during hand motor task (finger tapping) in a healthy subject with tDCS sponge electrodes placed over hand motor region. Clearly, the rectangular ‘deactivation’ seen as a green vertical line on the scalp (B), and that overlaps exactly with the position of the cathode, is an artifact (Shereen et al, in preparation).

Creating concurrent tDCS & fMRI acquisition and processing pipelines that consider, prevent, and correct for the above-mentioned potential pitfalls, and additional considerations, is an active area of research. Most likely, a combination of improvements in pulse sequence design, MRI-compatible tDCS hardware, and concurrent tDCS/fMRI specific field inhomogeneity correction techniques will need to be developed in concert to address these issues. It is interesting to note that the activation maps in Figure 9.B were created comparing time blocks of finger-tapping versus rest, *all during the tDCS condition*. Intuitively, any tDCS related artifacts would arise when comparing fMRI with tDCS on versus off, but would cancel when analyzing activations *within* the tDCS on condition. A possible explanation of this unintuitive artifact is that slight movements associated with finger tapping may be correlated with large signal intensity changes resulting from altering the magnetic field distribution associated with tDCS, particularly near the electrodes where it is strongest. Whatever the explanation, artifact removal is not trivial and may depend on specific tasks, imaging sequences, and processing methods. Ultimately, if artifacts are not obvious and direct artifact removal is not experimentally considered, then the possibility to misinterpret false positive fMRI activation as being caused by tDCS effects on neuronal activity is increased.

## 6. Gyri-precise finite element head models to address variability in MR imaging studies

A central challenge in optimizing tDCS efficacy is individual variability, which limits population-level effect size (Ammann, Lindquist, & Celnik, 2017; Lukasik et al., 2018; Wiethoff, Hamada, & Rothwell, 2014) and highlights the potential importance of dose customization (Kessler et al., 2013). Two main sources of variability are: 1) anatomical, leading to variation in brain current flow pattern and intensity among individuals (regional electric fields, EF;(Csifcsák, Boayue, Puonti, Thielscher, & Mittner, 2018; Datta, 2012; I. Laakso, Tanaka, Koyama, De Santis, & Hirata, 2015; Ilkka Laakso et al., 2016)), and 2) brain state, which leads to different responses even for the same brain current flow pattern (Chrysikou, Berryhill, Bikson, & Coslett, 2017; Esmaeilpour et al., 2018).

Computational models of current flow are well suited to address anatomical variability and can be retrospectively used as a regressor in tDCS-imaging data to control for differences in response based on individual brain current flow. Simulation of EF distribution based on high resolution structural images have informed study design and optimization of stimulation parameters in tDCS (Datta et al., 2009b) and model predictions have been repeatedly validated (Datta, Zhou, Su, Parra, & Bikson, 2013; Esmaeilpour et al., 2017; Y. Huang, A. A. Liu, et al., 2017; Opitz et al., 2016). The precise pattern of electric field distribution through the brain is determined by stimulation dose (i.e. electrode location and current intensity) as well as underlying anatomy and tissue properties (Marom Bikson et al., 2012). With the development of increasingly automated MRI-based individualized modeling pipelines (Huang, Datta, Bikson, & Parra, 2017), regression of current flow in imaging studies is increasingly accessible. However, the relationship between regional current flow intensity and neuromodulation remains unclear and may not be linear or even monotonic (Esmaeilpour et al., 2017). Conversely, imaging data can be used to retrospectively address the role of brain state in response variability. Combining current flow modeling and imaging, as well as behavioral and other biomarkers, can explain variability in how brain current flow and brain state influence and shape the effects of tDCS – but as we will explain, such hypothesis testing requires nuance.

Regional functional changes can be correlated to regional current intensity making the combination of current flow modeling and imaging in principle tractable (Halko et al., 2011). The regression can be done across the whole head (i.e. comparing the two maps of EF and fMRI data in the entire brain and connectivity values between different regions), as well as in specific regions of interest (ROI) that are segmented based on study hypothesis (i.e. comparing averaged EF and brain activity in each ROI). The anatomical images needed for individual models (Datta et al., 2009a) should have a wide field of view (all scalp down to neck) and high resolution (i.e. at least 1 mm) to support ideal current flow modeling (Y. Huang, A. A. Liu, et al., 2017).

Local EF intensities do not have a trivial relationship with resulting neuromodulation in neuronal network activity level and ultimately complex behaviors (Esmaeilpour et al., 2018). Therefore, combining imaging data and current flow predictions requires assumptions on 1) the scaling of “neuromodulation” with regional EF (which itself requires decision on if to use maximum e-field, average, or some other aggregate measure) and 2) the measure of brain “activation” based on imaging. Indeed, the sub-threshold nature of tDCS may make the effects of stimulation state dependent in a region-specific manner, but then regions may also interact. The result is a complicated mosaic of interactions that neither current flow or imaging in isolation can dissect.

However, if tDCS produces a strong enough EF to affect targeted areas of the brain, it would also affect several other brain regions simultaneously using conventional montages (i.e. large sponge electrodes). More to the point, brain regions do not operate in isolation, they rather communicate in micro, meso and large-scale networks (Polanía, Nitsche, & Paulus, 2012). Diffusivity of current flow with conventional tDCS application (Fig 10) in a complex, highly active and interconnected network, complicates intensity-response studies in a voxel-wise manner. Intensity-response depends on how a targeted brain region responds to the applied EF and how other brain areas interact with the targeted area. Its highly important to include connectivity analysis (i.e. functional and/or structural) in dose-response studies in tDCS-MR imaging.

**Figure 10:**
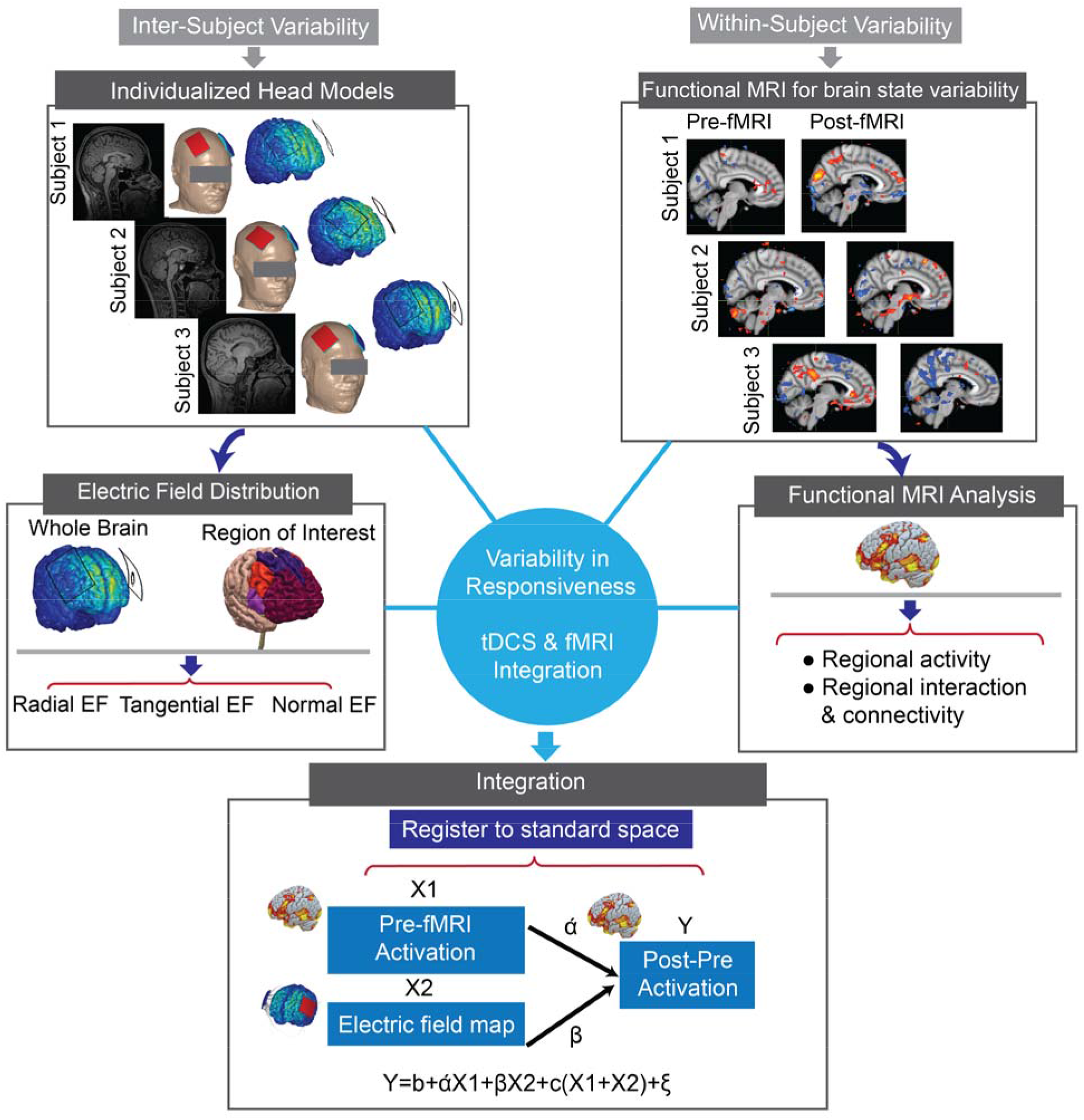
Workflow of computational modeling integration with functional MRI. Given the two separate maps of standard space used in fMRI and head models used in modelling, in the first step, high resolution EF map in fine meshed geometry should be interpolated to voxel-wise standard space. Then, both head models and functional images should be registered to standard space to enable comparison.

Technically, given the two maps of EF and brain activation, in the first step, the two sources of information should be in the same space with the same resolution (Fig 10). The next question is whether variability in EF intensity and ongoing brain activation can explain variability in the resulting change in neural activation and ultimately in the behavioral outcome. Based on the specific study hypothesis, cortical regions could be segmented using standard structural atlases allowing an assessment of the effect of EF and baseline ongoing activation in the given structural region. Alternatively, cortical areas could be segmented based on EF intensity distribution. Moreover, it could be informative to investigate the effect of EF direction (e.g. radial or tangential) on neural activation (Marom Bikson et al., 2004; Fischer et al., 2017b; Radman, Ramos, Brumberg, & Bikson, 2009).

### 6.1. Case study: technical issues for intensity-response questions

The relationship between applied current over skin and induced electric field in brain is linear and determined by the physics of tDCS (i.e. electric field intensity in the brain will increase linearly with applied current; more applied current, more electric field in brain (M. Bikson et al., 2015; Esmaeilpour et al., 2018)). However, the challenging question is whether increasing current intensity necessarily increases neural and behavioral responses? We do not have a clear understanding of intensity responses at the neural network level. In our case study, in a cross-over, sham controlled study in a population of methamphetamine abstinent subjects (as described in previous section, Fig 4 A), the main question was simply: is there a monotonic relationship between induced EF in each brain region and changes in neural activation using fMRI? Each participant was assigned to receive both active and sham arms in a randomized order and a conventional montage (bipolar balanced, bilateral DLPFC, 5×7 cm electrodes, 2 mA) was used for tDCS. To determine whether the EF induced in the targeted brain area could explain variability in fMRI response to tDCS (i.e. changes in activation after stimulation compared to before), prefrontal cortex (PFC) was segmented into different structural regions using FSL based on the Harvard Oxford atlas (Fig 11. A) and registered to MNI space of each subject (Smith et al., 2004). Regions of interest included separate masks for Frontal Pole (FP), Superior Frontal Gyrus (SFG), Medial Frontal Gyrus (MFG), Frontal Orbital Cortex (FOC) and Frontal Medial Cortex (FMC). Based on modeling results using a bilateral DLPFC montage in this study, majority of prefrontal cortex will receive electric field (Fig 11. B). Another method of segmentation could be based on current flow distribution profile. Regions of interest in current-flow segmentation in our study include: Superior Frontal Gyrus (SFG), Ventrolateral Prefrontal Cortex (vlPFC), medial Orbitofrontal Cortex (mOFC) and Frontal Pole (FP). Our results did not show any correlation in any of the ROIs in current-flow segmentation and atlas-based segmentation except Frontal Pole (r=0.7, *p*=0.0041). Statistical tests were corrected for multiple comparisons (Bonferroni correction). FP was the area that received maximum EF compared to other regions (Fig 11). However, although there was a correlation between current intensity and activation changes in FP, diffusivity and lack of clear targeting complicated the analysis of intensity-response in this case study. Regressing out interactive effects between different cortical regions is challenging when e-field modeling reveals that all frontal brain regions are functionally engaged by conventional montages. Using montages that produce more focal stimulation such as HD or using functional connectivity analyses that incorporate the interactions between different brain regions could potentially provide critical insights about intensity-response relationship in tDCS at the neural network level.

**Figure 11:**
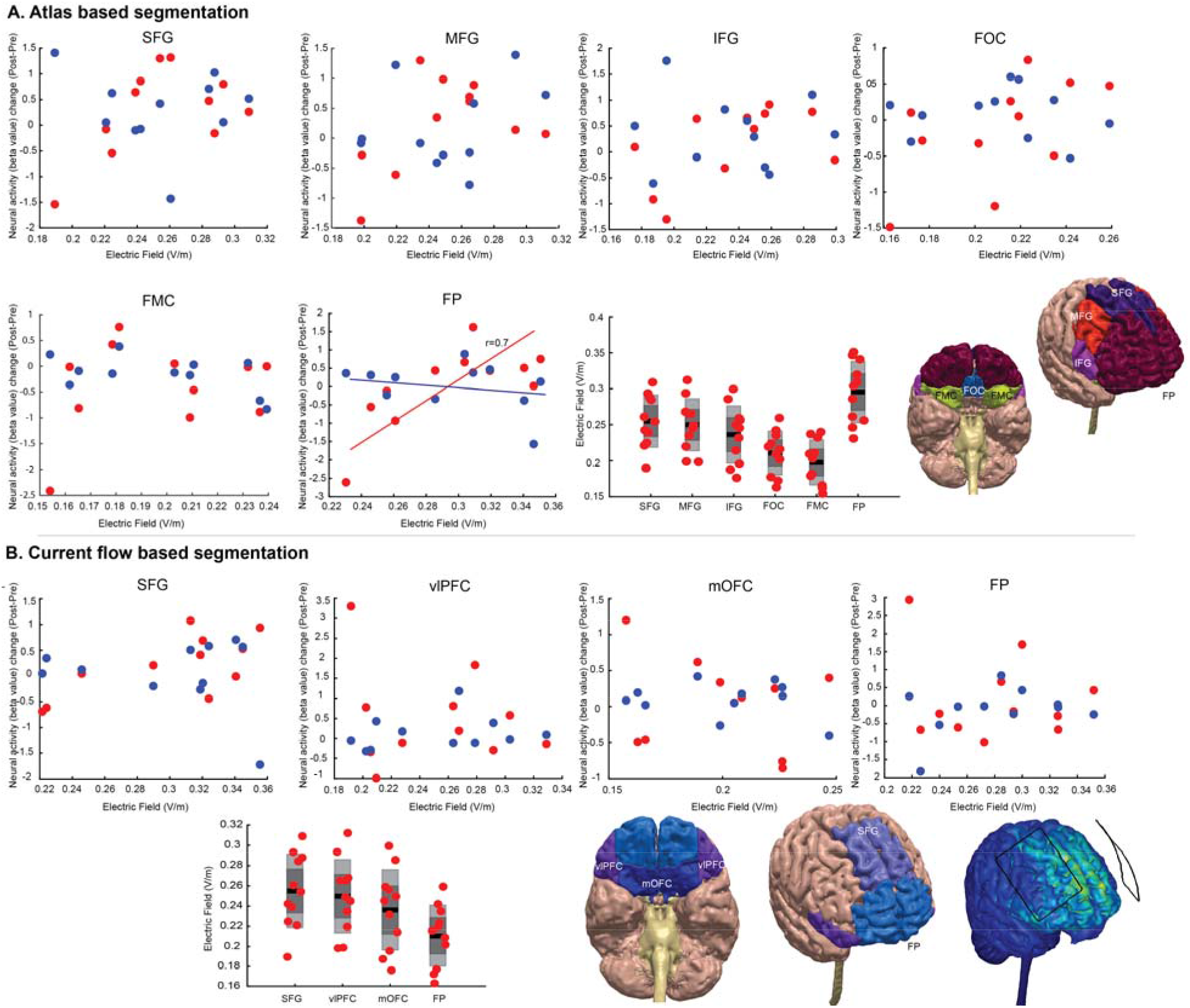
Correlation of electric field with neural activation change (post-pre) in the study introduced in figure 4. (A) Atlas based segmentation. Regions include: Frontal Pole (FP), Superior Frontal Gyrus (SFG), Medial Frontal Gyrus (MFG), Frontal Orbital Cortex (FOC) and Frontal Medial Cortex (FMC). (B) Current flow-based segmentation. Areas are selected based on current flow pattern generated based on bilateral DLPFC montage, 5×7 electrodes. Regions of interest include: Superior Frontal Gyrus (SFG), ventrolateral Prefrontal Cortex (vlPFC), medial Orbitofrontal Cortex (mOFC) and Frontal Pole (FP). Blue circles: sham stimulation, red circles: active stimulation.

## 7. Concluding remarks

This methodological review explains the parameter space in tDCS-MR imaging trial design. Functional MR imaging could contribute to answer predictive, mechanistic and localization questions regarding neural modulation resulting from tDCS stimulation. tDCS-MR imaging can potentially address important questions on functional correlates of tDCS mechanisms. However, to be meaningful, trial design requires nuanced understanding of both imaging and tDCS biophysics. For example, protocols optimized for either imaging or tDCS in isolation may not be suitable for combined tDCS-fMRI studies. Leveraging MR-based functional neuroimaging methods as indicators of neural states and traits for predicting variance in response to tDCS, requires carefully integrated study design. Also, individualized finite element head models provide a measure of anatomical variability in tDCS studies and could be utilized to regress out this variability in fMRI studies. In research studies with the intention of addressing dose-response relationship, use of conventional tDCS protocols with two large pads, as opposed to High-Definition (HD) tDCS, complicates analysis, especially because of how different brain regions interact functionally. Therefore, more focal stimulation protocols and/or using functional connectivity analyses that incorporate the interaction between brain regions along with induced electric field in each cortical area could be a better method to approach these questions in tDCS-MR imaging studies. We further discussed the importance of MR compatible devices and potential source of artifact that should be carefully investigated when imaging concurrently with stimulation.

## Data Availability

Data, list of studies include in the systematic review and/or code used in this study are available in the public domain upon direct request from authors.

## Acknowledgments

This work was supported (to MB) National Institutes of Health: NIH–NINDS 1R01NS101362, NIH–NIMH 1R01MH111896, NIH–NCI U54CA137788/U54CA132378, and NIH–NIMH 1R01MH109289. HE received supports from Warren K Family foundation and supported by a NARSAD young investigator grant (27305) from Brain and Behavior Research Foundation for this work. ZE was supported by Iranian Cognitive Sciences and Technologies Council.

